# Mpox detection in the wastewater and the number of hospitalized patients in Poznan, Poland

**DOI:** 10.1101/2022.12.23.22283918

**Authors:** Monika Gazecka, Jakub Sniezek, Krzysztof Maciolek, Arleta Kowala-Piaskowska, Pawel Zmora

## Abstract

We monitored for mpox in the Poznan wastewater system from July to December 2022 and compared its occurrence with the number of hospitalizations. Our results suggest that the scale of the epidemic is underestimated, and many mpox-infected individuals are not identified by the public health authority.

**Article summary line:** Wastewater-based epidemiology can determine the scale of an epidemic and estimate the number of infected individuals not under public health authorities.

---

The PCR-based quantitative and qualitative analysis of pathogens in wastewater is the principle of wastewater-based epidemiology (WBE), a useful tool for predicting the scale of epidemics and the genetic diversity of pathogens. WBE allows for the monitoring of the spread of pathogens in populations when traditional epidemiological approaches are limited due to low access to health services and diagnostic tests, overwhelmed diagnostic laboratory capabilities, society’s reluctance to diagnostic test, and the high availability of “do it yourself” rapid cassette tests (1,2). Unlike WBE, traditional approaches can miss cases of infected individuals who intentionally do not want to be tested due to the social stigma associated with some diseases, including mpox. WBE also allows for the tracing of asymptomatic infections and the cryptic transmission of pathogens (1–3). The effectiveness of WBE as an indicator of pathogen infection and its prevalence in the population has already been demonstrated for poliovirus, measles virus, enteroviruses, hepatitis A and E viruses, noroviruses, influenza A virus, dengue virus, and SARS-CoV-2 (1–3). More recently, mpox genetic material was detected in wastewater collected from five districts in the Netherlands (4), eight out of nine wastewater treatment plants (WTPs) in San Francisco, California, USA (5), five out of ten WTPs in Miami-Dade County, USA (6), WTPs from Paris, France (7), and airport buildings in Rome, Italy (8). However, to date, the detection of mpox in wastewater has not been correlated with the number of hospitalized infected individuals.

Mpox is endemic in Central and Western African countries, and outbreaks outside Africa have been associated with international travel and/or contact with imported animals. Since May 2022, increasing numbers of mpox cases have been reported in European and North American countries; therefore, on July 23, 2022, WHO declared mpox a public health emergency of international concern. A total of 82,999 cases of mpox and 66 deaths have been reported worldwide, including 214 cases in Poland as of December 19, 2022 (9). However, since the relatively mild course of mpox, these number may be highly underestimated.

Therefore, the aim of our study was to monitor for mpox virus (MPV) in wastewater from two WTPs in Poznan, Poland, and compare this with hospitalization numbers.

From July to December 2022 (weeks 27–48), we collected daily average wastewater samples twice a week (on Tuesdays and Thursdays) from the Central WTP (CWTP) and the Left Bank WTP (LWTP), with capacities of 200,000 m^3^/d and 50,000 m^3^/d, respectively. Total genetic material was isolated, and MPV was detected by commercially available qPCR TaqMan-specific probes and primers (Vi07922155_s1; ThermoFisher). Samples were considered positive if the cycle threshold was below the 40th cycle. The number of newly admitted mpox patients per week was obtained from the Department and Clinic of Infectious Diseases, Hepatology and Acquired Immunodeficiencies (DCIDHA), Poznan University of Medical Sciences. The DCIDHA is the only medical unit in Poznan that admits and reports MPV-infected individuals. According to Polish jurisdiction, mpox patients must be hospitalized and reported to public health authorities; therefore, the number of admitted patients should represent the scale of the epidemic.

MPV genetic material was detected in weeks 29, 43, and 47 at the CWTP and in weeks 37–38 and 40– 43 at the LWTP. A total of 22 mpox patients were admitted to the DCIDHA from July to December 2022, with the highest number of hospitalized individuals from mid-July to mid-August. The detection of MPV in week 29 at the CWTP may have been associated with the highest number of admitted patients a week earlier, since the DCIDHA wastewater is transported to this WTP. However, the detection of MPV at the LWTP from mid-September to mid-October did not correlate with the number of reported mpox cases. This suggested a relatively high number of non-identified MPV-infected individuals and a local epidemic foci in the Poznan population.

The ongoing mpox epidemic is caused by the West African clade MPV, which causes mostly mild symptoms (i.e., fever, rash, and swollen lymph nodes). However, the currently circulating strain of MPV can cause also atypical symptoms, such as the presentation of only a few or even just a single lesion, lesions limited to the genital or perineal/perianal area, or no lesions at all (9). Mpox patients with these mild symptoms may not seek clinical treatment and are thus not reported. In addition, some individuals may deliberately avoid the health care system due to the social stigma surrounding mpox. Leading to the non-reporting of mpox, these factors may bias official reports of the scale of the epidemic (9). Therefore, WBE is a promising additional tool that can complete data gathered by the clinical monitoring approach and predict more accurately the development and progress of the current mpox outbreak.

**Figure 1.**
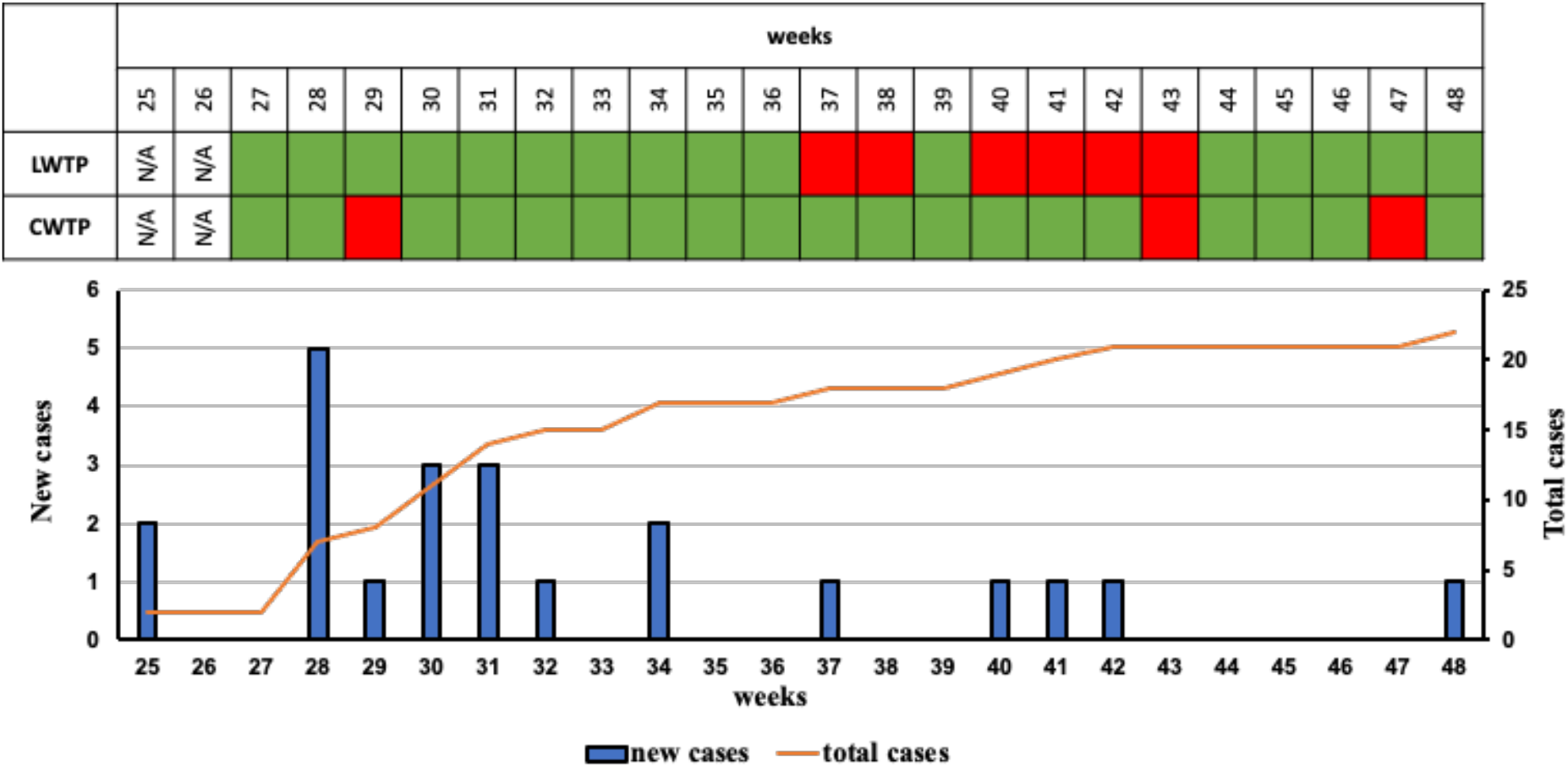
Mpox virus detection in the Poznan wastewater treatment plants and number of mpox patients hospitalized at the Department and Clinic of Infectious Diseases, Hepatology and Acquired Immunodeficiencies, Poznan University of Medical Sciences. Red indicates a positive sample, green indicates a negative sample, N/A – not analyzed. LWTP – Left Bank wastewater treatment plant, CWTP – Central wastewater treatment plant.

## Data Availability

All data produced in the present work are contained in the manuscript

## Acknowledgements

The project was partially funded by the Municipal Economy Department, City of Poznan, and statutory funds of the Institute of Bioorganic Chemistry Polish Academy of Sciences and Aquanet Laboratorium. We would like to thank the Sampling Laboratory, Aquanet Laboratorium for their technical help with the samples collection.

## About the Author

Monika Gazecka, M.Sc. is a researcher at the Department of Molecular Virology, Institute of Bioorganic Chemistry Polish Academy of Sciences. Her primary interest focus on the emerging viruses, wastewater-based epidemiology, molecular epidemiology and virus-host interactions.

